# Comparative Analysis of Three Surveys on Primary Care Providers’ Experiences with Interoperability and Electronic Health Records

**DOI:** 10.1101/2024.01.02.24300713

**Authors:** Nathaniel Hendrix, Natalya Maisel, Jordan Everson, Vaishali Patel, Andrew Bazemore, Lisa S. Rotenstein, A Jay Holmgren, Alex H. Krist, Julia Adler-Milstein, Robert L. Phillips

## Abstract

**Objective:** This study compared primary care physicians’ self-reported experiences with Electronic Health Records’ (EHR) interoperability, as reported across three surveys: the 2022 Continuous Certification Questionnaire (CCQ) from the American Board of Family Medicine, the 2022 University of California San Francisco’s (UCSF) Physician Health IT Survey, and the 2021 National Electronic Health Records Survey (NEHRS).

**Materials and Methods:** We used descriptive analyses to identify differences between survey pairs. To account for weighting in NEHRS and UCSF, we assessed the significance of differences using the Rao-Scott corrected chi-square test.

**Results:** CCQ received 3,991 responses, UCSF received 1,375 from primary care physicians, and NEHRS received 858 responses from primary care physicians. Response rates were 100%, 3.6%, and 18.2%, respectively. Substantial and largely statistically significant differences in response were detected across the three surveys. For instance, 22.2% of CCQ respondents said it was very easy to document care in their EHR, compared to 15.2% in NEHRS, and 14.8% in the UCSF survey. Approximately one-third of respondents across surveys said documenting care in their EHR was somewhat or very difficult. The surveys captured different respondent types with CCQ respondents trending younger, and NEHRS respondents more likely to be in private practice.

**Discussion:** All surveys pointed to room for improvement in EHR usability and interoperability. The differences observed, likely driven by differences in survey methodology and response bias, were likely substantial enough to impact policy decisions.

**Conclusion:** Diversified data sources, such as those from specialty boards, may aid in capturing physicians’ experiences with EHRs and interoperability.

## Introduction

The United States Centers for Disease Control and Prevention began conducting the National Electronic Health Records Survey (NEHRS) with funding from the Office of the National Coordinator for Health IT in 2012 with the goal of reporting on national rates of electronic health record (EHR) adoption and patterns of EHR use across office-based physicians in the United States.(1) It has been conducted almost annually ever since. Researchers and policymakers have used NEHRS to evaluate and understand how changes in EHR adoption and other healthcare information technologies (HIT) impact clinical practice. More recently, policymakers have used NEHRS to examine office-based physicians’ engagement in electronic exchange of health information and interoperability of EHR systems.(2) Assessing these HIT functions are important to the Medicare Promoting Interoperability Program and the strategic objectives of the 21^st^ Century Cures Act and actively used by policymakers.(3)

Despite the important information it has provided about clinicians’ transition from paper to electronic records, to date, NEHRS has not been statistically compared to other sources of information on physician EHR use. Surveys of physicians and other professions are challenging and often feature low response rates, raising the possibility of considerable response bias. Response rates in recent years of NEHRS have been as low as 25%, perhaps reflecting well-known challenges in recruiting healthcare professionals to complete surveys.(4,5) Researchers analyzing NEHRS do not have the information required to understand the importance of missing information due to a low response rate and potentially non-representative response cohort. It is unclear whether these are actual or merely theoretical limitations of NEHRS.

In this study, we compared responses to three surveys, each intended to gather information on physicians’ use of EHRs but fielded with substantially different strategies: 1) the 2021 NEHRS; 2) the 2022 Continuous Certification Questionnaire (CCQ) from the American Board of Family Medicine (ABFM); and 3) the inaugural version of University of California, San Francisco’s (UCSF) Physician Health IT Survey, which was also fielded in 2022. The NEHRS was a voluntary survey fielded among a relatively small sample with repeated mailing and active searches to identify the address and contact information for targeted respondents. The CCQ was a required component of Family Medicine physicians’ recertification process through ABFM. The UCSF Physician Health IT Survey was a voluntary survey designed to maximize number of responses but not response rates.

In comparing responses, we focused on the characteristics of responding physicians and the domain of interoperability among physicians providing primary care, as this topic and these respondents were common across all three surveys and assessed using comparable questions. We sought to identify trends common to all three surveys, but as importantly, discrepancies that may highlight potential paths for improved data collection. Policies affecting EHR use and implementation aim to improve interoperability which is vital to patient care. Understanding the strengths and limitations of different methods for ascertaining these policies’ impacts on physicians is critical to inform policymakers, regulators, EHR vendors, and health system leaders working to improve various dimensions of EHR functionality.

## Methods

### Population and Instruments

We compared three surveys covering physicians’ experiences with interoperability in EHRs: the 2021 NEHRS, the 2022 CCQ, and the 2022 UCSF Physician IT Survey (Table 1).

**Table 1:** Comparison of survey processes.

|  | **ABFM CCQ** | **UCSF Physician Health IT Survey** | **NEHRS** |
| --- | --- | --- | --- |
| Number of respondents | 3,991 | 1,375 primary care; 3209 total | 858 primary care; 1,875 total |
| Response rate | 100% | 3.6% | 18.2%* |
| Survey Frame | ABFM Diplomates | Physicians with a listed email in Definitive Healthcare data | American Medical Association and American Osteopathy Association Master files |
| Breadth of Specialties | Family Medicine Physicians | Random sample of all specialties providing direct patient care | Stratified sample of all specialties providing direct patient care |
| Strategies to garner response rate | Required for continuation of certification | Repeated contact; 8 emails, 0-2 mailings, 0-1 postcard depending on condition | Repeated contact (7 emails, 4 mailings and 1 postcard) and tracing—manually identifying contact information. |
| Survey length and burden | 2.5-page survey per respondent (participants randomized to modules) | 6-page survey | 4-page survey |

ABFM CCQ = American Board of Family Medicine Continuous Certification Questionnaire; NEHRS = National Electronic Health Record Survey; UCSF = University of California, San Francisco. \*This differs from the response rate reported in NEHRS public documentation and reflects the number of completed responses by eligible respondents divided by the total number contacted, excluding physicians who were identified as ineligible.

Physician informants asked to complete the NEHRS were selected through a random recruitment process from the American Medical Association and American Osteopathy Association Master Files, stratified by specialty and region.(4) They were initially sent both mail and electronic recruitment letters and, subsequently, mail and electronic versions of the survey. The survey included questions intended to assess eligibility for participation: specifically, participants were required to spend most of their working time providing patient care, not be federally employed, and be less than 85 years of age at the time of the survey. Although NEHRS samples physicians in primary care, specialty care, and surgery, we included only primary care physicians in this analysis and excluded any respondent who said that they do not use an EHR. In the NEHRS public use file, each individual respondent was assigned a sample weight based on region and specialty. These weights were essential to accurately interpreting NEHRS data and were accordingly integrated into the analysis.

Completion of the CCQ has been required of the more than 102,000 family physicians’ participating in continuous certification processes since 2017.(6) As such, it has a 100% response rate. For nearly a decade the ABFM has included questions about EHR Meaningful Use policies and, for the 2022 CCQ, ABFM collaborated with the United States Office of the National Coordinator for Health Information Technology to incorporate questions about EHR use that closely parallel those in NEHRS. The goal of this collaboration was to enable comparison and to evaluate the potential utility of supplementing NEHRS with outside data. All respondents to the CCQ first answered a set of personal, practice, and demographic questions before being randomized to one of two modules on EHR usage, including one on interoperability. Finally, they were randomized to one of five modules that covered topics such as burnout and meaningful use of EHRs. Thus, EHR module cohorts were 50% of the overall number of respondents, and cohorts for other modules were 20%. Only respondents who indicated that they use an EHR system and that they provided direct patient care were included in the analysis. To ensure comparability with NEHRS, we also excluded federally employed physicians.

The UCSF Physician Health IT Survey was initiated in 2022 to collect in-depth information on how information technology is integrated into clinical settings. Researchers used simple random sampling to select 90,000 potential participants with listed email addresses from Definitive Healthcare, a proprietary dataset designed for healthcare analytics. All sampled physicians received 8 emails. 60,000 sampled physicians also received a postcard reminder to complete the survey. 30,000 sampled physicians also received 2 recruitment letters by mail. The UCSF survey recruited physicians of all specialties, but only physicians who used EHRs and who worked in primary care for non-federal employers were included in this analysis.

### Statistical analysis

Both NEHRS and the UCSF survey used survey weights with the goal of improving the generalizability of their findings. We therefore used a Rao-Scott corrected chi-square test, which adjusts the chi-square estimate for weights to account for changes in the composition of the population.(7) Since our interests were in identifying specific differences in the surveys, we conducted significance tests in a pairwise fashion across the three surveys.

While the ABFM CCQ and the UCSF survey both based questions on NEHRS, some questions were phrased differently or included different answer options. We did not statistically analyze differences in the responses to questions that we found to be incomparable across surveys, but we did retain them in the tables for reference.

As a supplemental analysis, we also conducted stratified comparisons of especially important indicators of interoperability experience across age groups (less than 50 or 50-plus years), EHR platform (Epic or other), and practice type (private practice or other).

All analyses were conducted in R version 4.1.2, with the Rao-Scott corrected chi-square test implemented in the “survey” package version 4.1.1.(8,9)

### Data availability

The NEHRS data used in this research are publicly available through the website of the United States Centers for Disease Control and Prevention.(10) ABFM CCQ data may be accessed for IRB-approved projects subject to the approval of the ABFM Research Governance Board; please contact the corresponding author for details. The UCSF survey data underlying this article cannot be shared due to privacy restrictions.

## Results

### Response Rate Comparison

The CCQ had the highest number of respondents and response rate across the three surveys. A total of 3,991 respondents to the 2022 CCQ provided direct patient care and were otherwise eligible for inclusion in the analysis (100% response rate). Of the 10,302 physicians sampled for NEHRS in 2021, 1,875 were eligible and completed the survey (18.2% response rate); although we could not determine how many family physicians were in the NEHRS sample, 858 (48.7% of respondents) listed primary care as their specialty.(4) Among the 90,000 physicians sampled for the UCSF Physician Health IT Survey, 3,209 responded (3.6% response rate), among whom 1,375 (42.8% of respondents) practiced in primary care.

### Respondent Demographics

Respondents to the three surveys represented different physician demographics (Table 2). Respondents to the CCQ were the youngest: the share of respondents aged 35 to 44 (42.1%) was over twice that seen in the NEHRS (17.0%) and the UCSF survey (15.2%). Male and female genders were approximately equally represented in all surveys, although women outnumbered men (51.7% to 48.3%) in the UCSF survey.

**Table 2:**
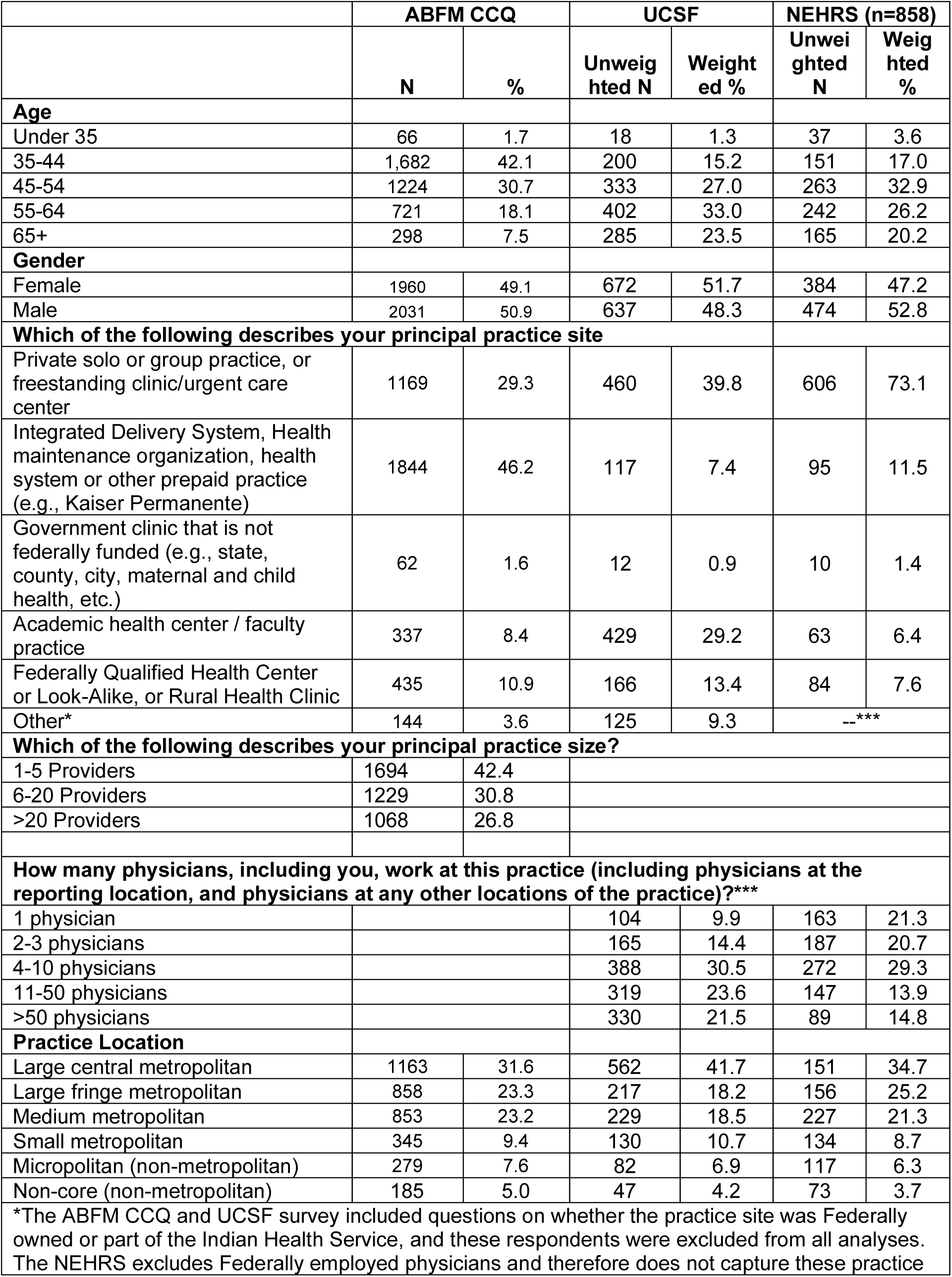
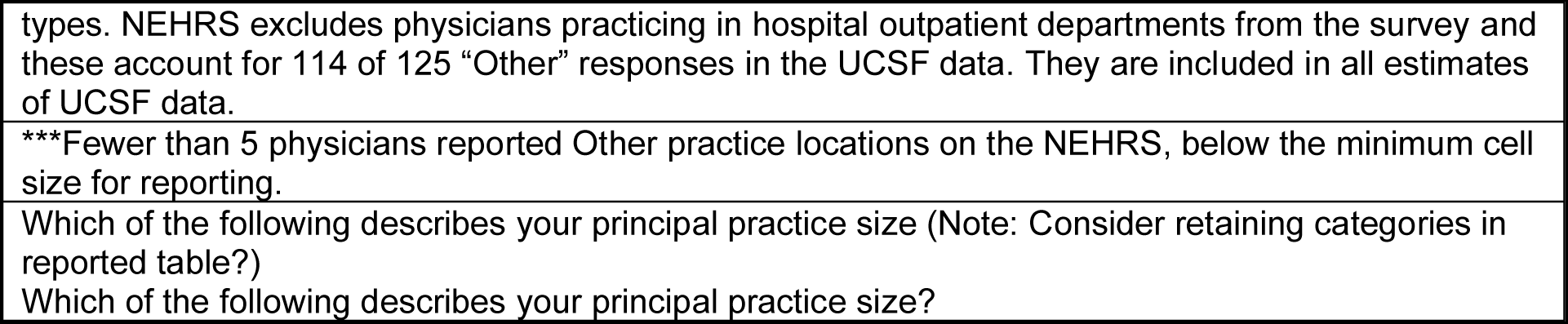
Survey respondent and practice characteristics.

|  | ABFM CCQ |  | UCSF |  | NEHRS (n=858) |  |
| --- | --- | --- | --- | --- | --- | --- |
|  | N | % | Unweighted N | Weighted % | Unweighted N | Weighted % |
| **Age** |  |  |  |  |  |  |
| Under 35 | 66 | 1.7 | 18 | 1.3 | 37 | 3.6 |
| 35-44 | 1,682 | 42.1 | 200 | 15.2 | 151 | 17.0 |
| 45-54 | 1224 | 30.7 | 333 | 27.0 | 263 | 32.9 |
| 55-64 | 721 | 18.1 | 402 | 33.0 | 242 | 26.2 |
| 65+ | 298 | 7.5 | 285 | 23.5 | 165 | 20.2 |
| **Gender** |  |  |  |  |  |  |
| Female | 1960 | 49.1 | 672 | 51.7 | 384 | 47.2 |
| Male | 2031 | 50.9 | 637 | 48.3 | 474 | 52.8 |
| **Which of the following describes your principal practice site** |  |  |  |  |  |  |
| Private solo or group practice, or freestanding clinic/urgent care center | 1169 | 29.3 | 460 | 39.8 | 606 | 73.1 |
| Integrated Delivery System, Health maintenance organization, health system or other prepaid practice (e.g., Kaiser Permanente) | 1844 | 46.2 | 117 | 7.4 | 95 | 11.5 |
| Government clinic that is not federally funded (e.g., state, county, city, maternal and child health, etc.) | 62 | 1.6 | 12 | 0.9 | 10 | 1.4 |
| Academic health center / faculty practice | 337 | 8.4 | 429 | 29.2 | 63 | 6.4 |
| Federally Qualified Health Center or Look-Alike, or Rural Health Clinic | 435 | 10.9 | 166 | 13.4 | 84 | 7.6 |
| Other* | 144 | 3.6 | 125 | 9.3 | --*** |  |
| **Which of the following describes your principal practice size?** |  |  |  |  |  |  |
| 1-5 Providers | 1694 | 42.4 |  |  |  |  |
| 6-20 Providers | 1229 | 30.8 |  |  |  |  |
| >20 Providers | 1068 | 26.8 |  |  |  |  |
| **How many physicians, including you, work at this practice (including physicians at the reporting location, and physicians at any other locations of the practice)?\*\*\*** |  |  |  |  |  |  |
| 1 physician |  |  | 104 | 9.9 | 163 | 21.3 |
| 2-3 physicians |  |  | 165 | 14.4 | 187 | 20.7 |
| 4-10 physicians |  |  | 388 | 30.5 | 272 | 29.3 |
| 11-50 physicians |  |  | 319 | 23.6 | 147 | 13.9 |
| >50 physicians |  |  | 330 | 21.5 | 89 | 14.8 |
| **Practice Location** |  |  |  |  |  |  |
| Large central metropolitan | 1163 | 31.6 | 562 | 41.7 | 151 | 34.7 |
| Large fringe metropolitan | 858 | 23.3 | 217 | 18.2 | 156 | 25.2 |
| Medium metropolitan | 853 | 23.2 | 229 | 18.5 | 227 | 21.3 |
| Small metropolitan | 345 | 9.4 | 130 | 10.7 | 134 | 8.7 |
| Micropolitan (non-metropolitan) | 279 | 7.6 | 82 | 6.9 | 117 | 6.3 |
| Non-core (non-metropolitan) | 185 | 5.0 | 47 | 4.2 | 73 | 3.7 |
| *The ABFM CCQ and UCSF survey included questions on whether the practice site was Federally owned or part of the Indian Health Service, and these respondents were excluded from all analyses. The NEHRS excludes Federally employed physicians and therefore does not capture these practice |  |  |  |  |  |  |

types. NEHRS excludes physicians practicing in hospital outpatient departments from the survey and these account for 114 of 125 “Other” responses in the UCSF data. They are included in all estimates of UCSF data.
\*\*\*Fewer than 5 physicians reported Other practice locations on the NEHRS, below the minimum cell size for reporting.
Which of the following describes your principal practice size (Note: Consider retaining categories in reported table?)
Which of the following describes your principal practice size?

The share of responses by setting varied across the three surveys. A disproportionate share of NEHRS respondents worked in private practice (73.1%) compared to CCQ (39.8%) or UCSF (29.3%). While a plurality of CCQ respondents were from health systems (46.2%), this setting was not well represented in either the USCF (7.4%) or NEHRS (11.5%) surveys. Far more respondents to the UCSF survey – 29.2% – practiced in academic health centers or faculty practices.

There was also some variability by practice size. Physicians from practices with more than 10 physicians were overrepresented in the UCSF survey (45.1%) versus NEHRS (28.7%). The CCQ asked about the number of providers working at the respondent’s main practice site instead of physicians at all locations of the practice, which is how NEHRS and UCSF surveys asked the question. Despite this more inclusive clinician language, a larger proportion of CCQ physicians (42.4%) indicated that they work in practices with 1 to 5 providers. Geographic representation was similar in the CCQ and NEHRS, but respondents to the UCSF survey were more likely to practice in central metropolitan locations compared to the other surveys’ respondents.

### EHR Vendor

There were significant differences in the EHR platform used by respondents and their satisfaction with these EHRs across the surveys (Table 3). While Epic was the most common EHR platform in all three surveys, Cerner was the second most commonly used platform in the UCSF survey and eClinical Works in the others. The CCQ had the largest share of respondents indicating that they do not know what EHR they use (1.8%). Respondents to NEHRS reported being very satisfied with their EHR at significantly higher rates (29.1%) than in the CCQ (26.7%) or UCSF survey (19.4%).

**Table 3:** Electronic health record (EHR) platform and user experience.

|  | ABFM CCQ |  | UCSF |  | NEHRS |  |
| --- | --- | --- | --- | --- | --- | --- |
|  | N | % | Unweighted N | Weighted % | Unweighted N | Weighted % |
| *EHR vendor\** |  |  |  |  |  |  |
| Allscripts | 211 | 5.4 | 50 | 5.3 | 48 | 5.0 |
| athenahealth | 372 | 9.6 | 75 | 8.1 | 94 | 9.7 |
| Cerner | 288 | 7.4 | 122 | 12.6 | 66 | 5.4 |
| eClinical Works | 431 | 11.1 | 87 | 9.1 | 110 | 13.9 |
| e-MDs | 32 | 0.8 | 8 | 0.8 | 16 | 2.2 |
| Epic | 1576 | 40.6 | 743 | 41.4 | 233 | 29.5 |
| NextGen | 172 | 4.4 | 47 | 4.8 | 46 | 7.2 |
| Practice Fusion | 84 | 2.2 | 9 | 0.9 | 21 | 3.1 |
| Greenway | 66 | 1.7 | 22 | 2.2 | 36 | 3.5 |
| Other | 582 | 15.0 | 139 | 14.2 | 183 | 20.2 |
| Don't know | 68 | 1.8 | 6 | 0.6 | 2 | 0.2 |
| *EHR satisfaction\*\** |  |  |  |  |  |  |
| Very satisfied | 1028 | 26.7 | 276 | 19.4 | 229 | 29.1 |
| Somewhat satisfied | 1496 | 38.9 | 480 | 35.3 | 336 | 39.1 |
| Neither satisfied nor unsatisfied | 314 | 8.2 | 114 | 9.1 | 77 | 7.4 |
| Somewhat dissatisfied | 630 | 16.4 | 281 | 22.6 | 130 | 15.2 |
| Very dissatisfied | 377 | 9.8 | 158 | 13.5 | 80 | 9.3 |
| *Ease of documenting care (ABFM CCQ and UCSF survey wording in parentheses)\** |  |  |  |  |  |  |
| Very easy (Excellent) | 420 | 22.2 | 202 | 14.8 | 134 | 15.2 |
| Somewhat easy (Good) | 882 | 46.5 | 546 | 40.4 | 402 | 47.3 |
| Somewhat difficult (Fair) | 475 | 25.1 | 429 | 33.6 | 265 | 33.7 |
| Very difficult (Poor) | 119 | 6.3 | 131 | 11.1 | 52 | 3.8 |

\* *p*-values for all comparisons < 0.001; \*\**p*-values for ABFM versus UCSF and NEHRS versus UCSF < 0.001, *p*-value for ABFM versus NEHRS = 0.875.

### EHR Use and Interoperability

Similarly, documentation and ease of documentation differed (Figure 1, Supplementary Material I). CCQ respondents reported spending more than 4 hours in after-work documentation at significantly higher rates (17.1%) than respondents in the other surveys (12.2% for UCSF and 8.4% for NEHRS). Interestingly, more CCQ respondents said that their experience of documenting care in the EHR was excellent / very easy (22.2%) versus the others (14.8% for UCSF, 15.2% for NEHRS).

**Figure 1.**
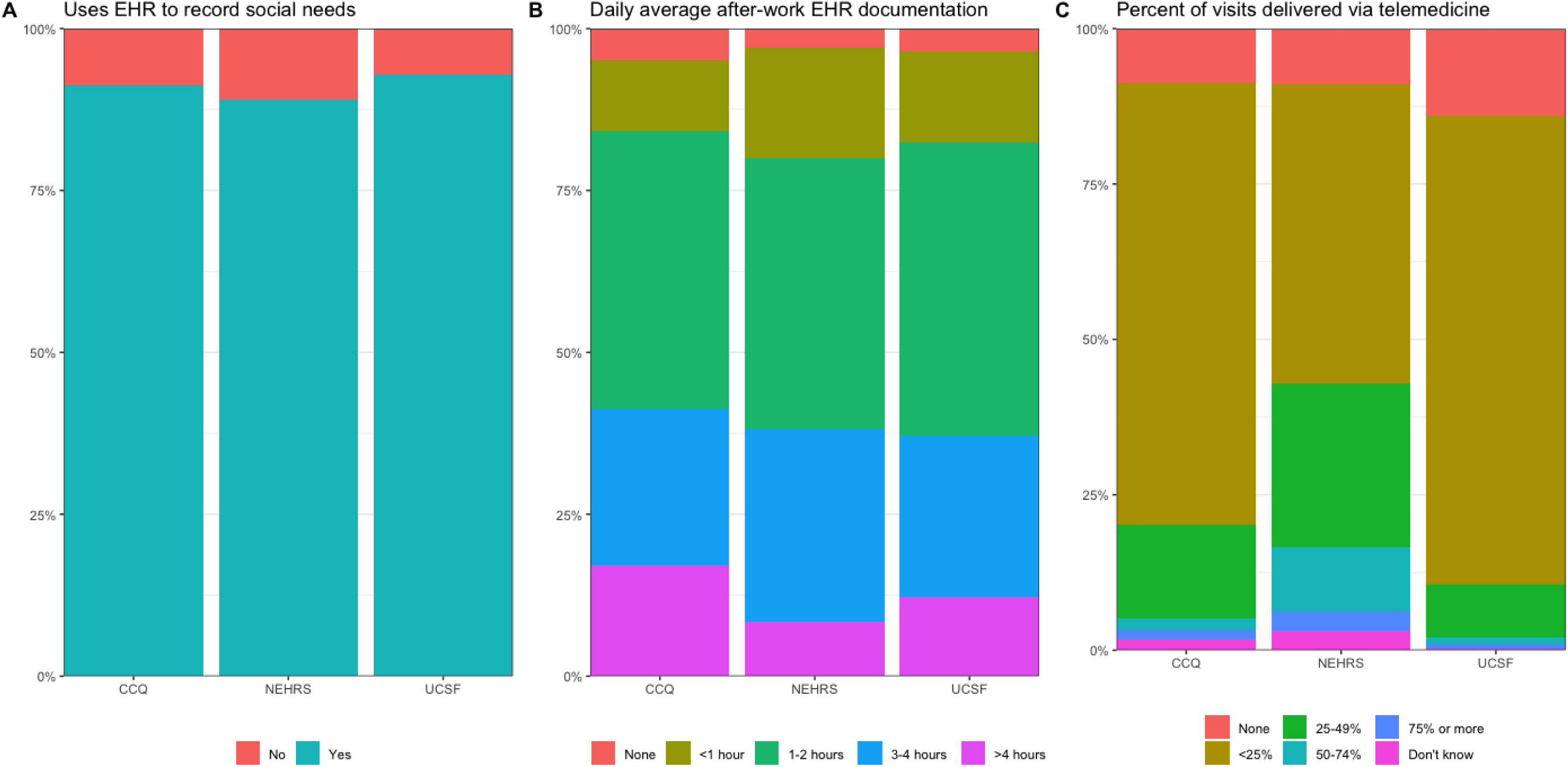
Comparison of documentation and practice patterns across the three surveys. For C, note that the CCQ and UCSF survey ask about telemedicine in the prior 3 months, while NEHRS asks about telemedicine since March 2020.

Several questions related to interoperability were similar across all three surveys; however, response options were identical for only two questions assessing two items on physicians’ experience with interoperability (Table 4). 45.8% of respondents to the CCQ and 45.4% of respondents to NEHRS indicated that their EHRs integrate patient health information from outside organizations into their EHR. 22.8% of respondents in CCQ and 21.9% of respondents to NEHRS indicated that they often had access to clinical information from outside organizations in their EHR. On both items, respondents to the UCSF survey were substantially less positive, with 37.6% of UCSF respondents indicating that their EHR integrated information and 14.6% indicating that they often had access to clinical information.

**Table 4:**
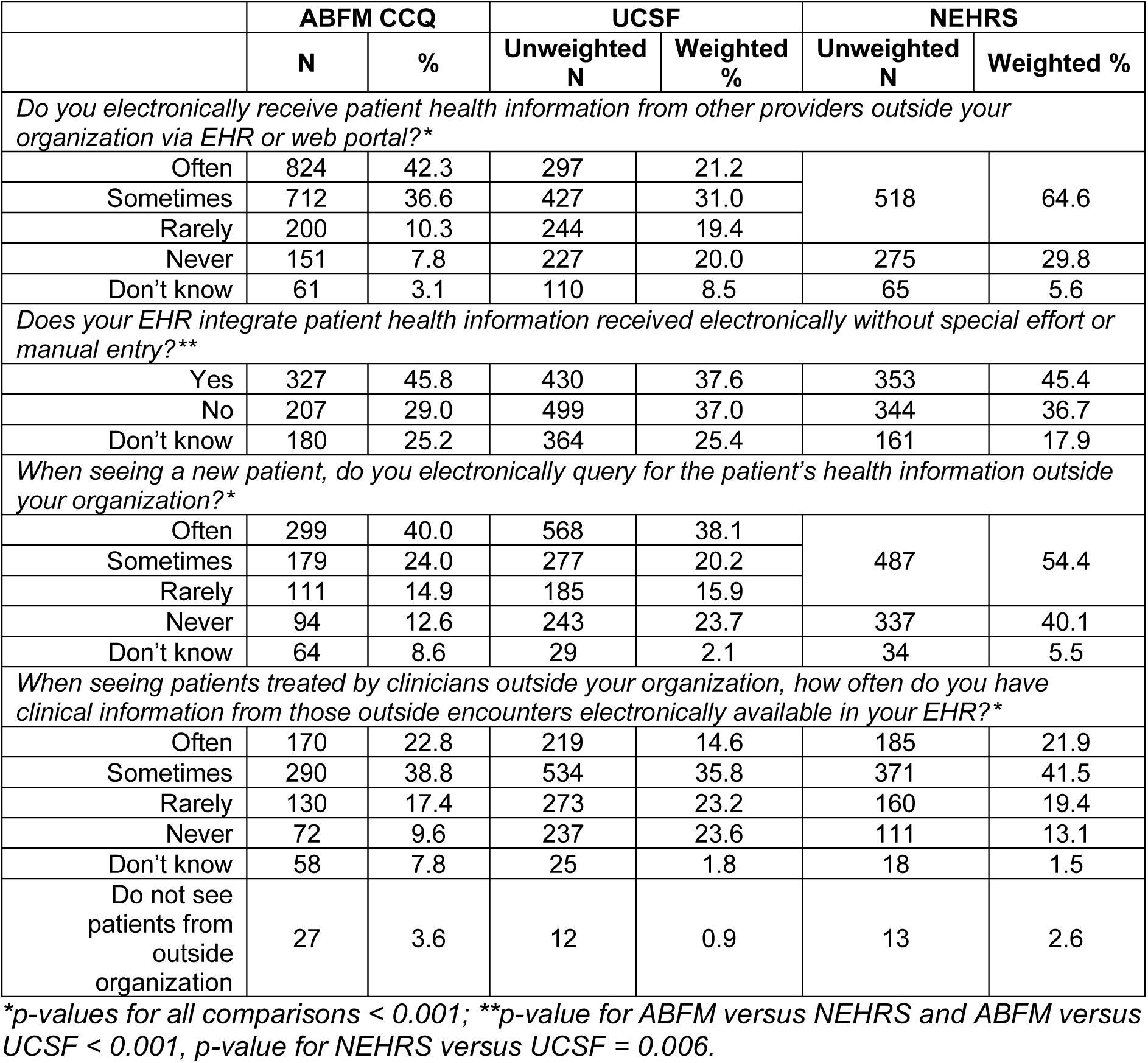
Interoperability experience.

Response options differed between NEHRs and the CCQ and UCSF survey for two additional interoperability options, making direct comparison across the three more challenging. For one of these items, respondents to the CCQ had a better experience of interoperability than respondents to the UCSF survey: 42.3% of CCQ respondents said that they often receive external information via EHR or web portal, compared to 21.2% of UCSF respondents. 64.6% of NEHRS respondents said that they received information but were not asked how often. For the second item, responses to the CCQ and UCSF were more similar: 40.0% of respondents to the CCQ indicated that they often queried for information from outside organizations for new patients compared to 38.1% of respondents to the UCSF survey. 54.4% of respondents to the NEHRS indicated that they queried for information, but again were not asked how often.

Some additional questions about the ease of using interoperable data were only asked in the CCQ and UCSF survey (Supplemental Material II). As before, CCQ respondents report better experiences with interoperability than the UCSF respondents do. For example, 24.1% of CCQ respondents say it is very easy to include external information in care decisions compared to 18.6% of UCSF respondents.

### Stratified Comparisons Across Surveys

We designed our stratified analysis to identify whether differences in survey composition — such as a greater number of CCQ respondents in the youngest age group — explained observed differences in opinions or if these differences persisted within sub-groups (Figure 2, Supplemental Material III). Results indicated that survey differences persisted across sub-groups: for instance, both older and younger respondents to the CCQ were more likely to say that information was integrated into their EHR than were the same age group respondents to the UCSF survey.

**Figure 2:**
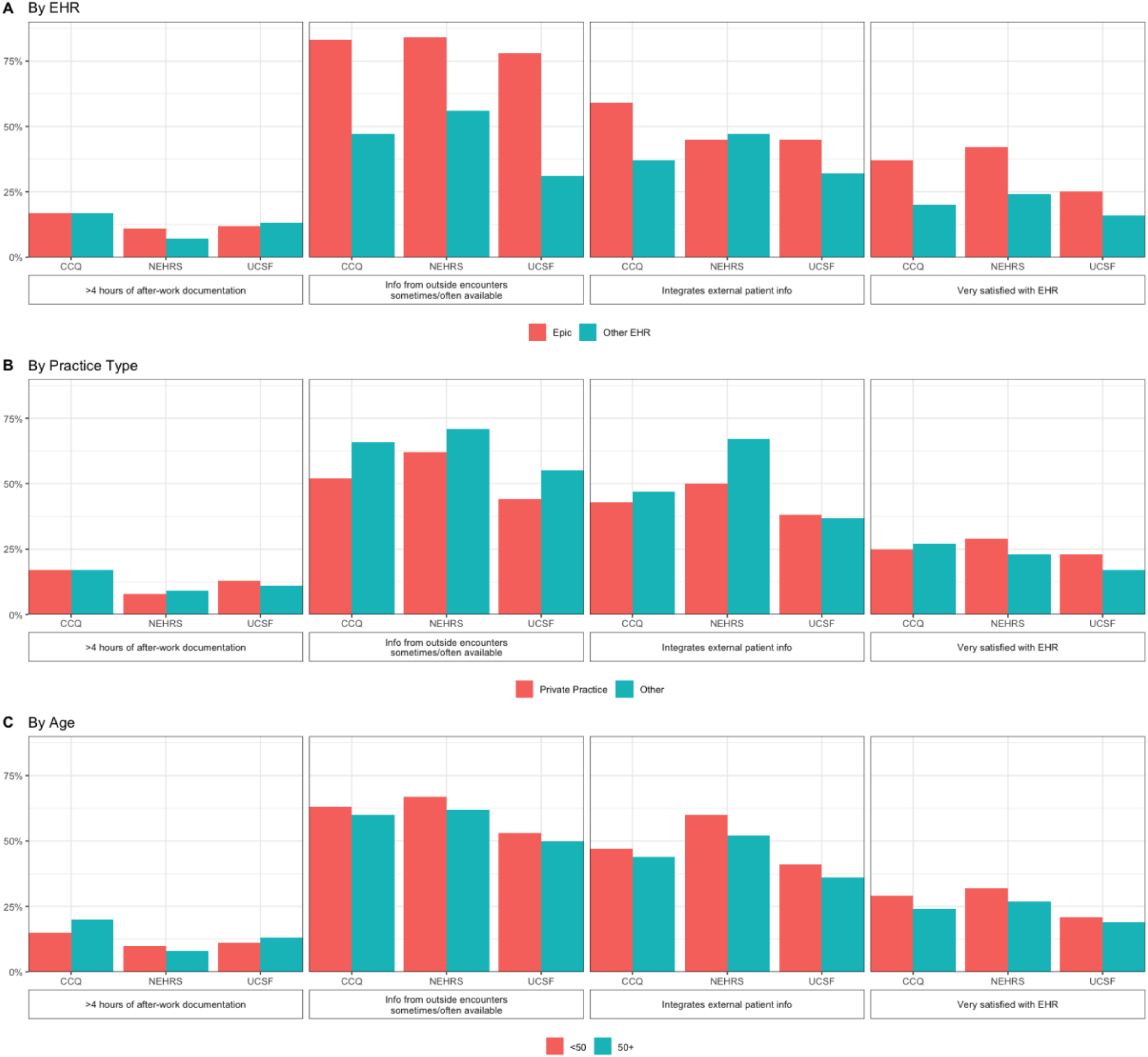
Stratified comparisons of key interoperability and EHR satisfaction measures

Differences between sub-groups were directionally consistent across most comparisons in the three surveys. For instance, all three found that Epic users were more likely to be very satisfied with their EHR, that physicians in private practice were less likely to have external patient health information available, and that physicians aged 50 years or more were less likely to integrate external information into care decisions.

## Discussion

These findings reflect a rare opportunity to compare primary care physician responses related to the aims of EHR policies across multiple surveys with different sampling strategies. The comparison of these three surveys expands our understanding of their validity, reliability, and generalizability. It also aids in determining whether improvements can be made to the current process of collecting information about interoperability and other dimensions of clinician experience with HIT. All three surveys revealed substantial opportunity for improvement in physician experience with HIT and particularly with interoperability. At the same time, the surveys responses significantly differed in several ways, suggesting that policymakers may benefit from integrating sources of information outside of NEHRS.

The surveys were unanimous in showing that physicians were not highly satisfied with EHRs, and that interoperability remains a significant problem. Approximately one quarter of physicians in all surveys indicated that they were very satisfied with their EHR platform. Similarly, approximately three-quarters of respondents reported documenting care for an average of one or more hours after work per day. Across surveys, only about 20% of physicians reported that information was often available from their patients’ healthcare encounters outside the primary care clinic. The three surveys indicated too that users of Epic were more likely to have this information available and that physicians working in private practice were less likely to.

Despite these common findings, almost all questions had statistically significant differences in the responses to the surveys. This suggests that the results of regression-based analyses would differ based on which of these data sources the researchers used, which may have substantial policy impacts. The observed differences likely arose in part from differences in respondents. The ABFM CCQ is a cross-sectional census with 100% response rate but from a single, large specialty. The plurality of CCQ respondents were aged 35 to 44, younger than in the other surveys. They were also over four times as likely to work at a health system compared to UCSF and NEHRS respondents. NEHRS may also have sampled from or had a response bias favoring solo practices and small clinics at a higher rate than the other surveys. However, substantial differences persisted in cross-survey comparisons among physicians who seemed similar.

Response bias likely played an important role in these observed differences, as the UCSF survey and NEHRS had much lower response rates than the CCQ with its 100% response rate. Some of the differences point towards the CCQ’s unique capture of data from respondents whose experiences are especially important to decisions in HIT policy. The response rate comparisons show that physicians are less likely to respond to discretionary, voluntary surveys such as NEHRS and UCSF, potentially making them less representative of the physician population at large. Our findings show that differing survey composition has downstream effects on the results and also indicate that there are likely differences across surveys not accounted for by observable differences in the composition of respondents. At the same time, the CCQ’s response rate, achieved by tying survey completion to the recertification process, may have a downside inasmuch as respondents may be more motivated to provide quick rather than thoroughly considered answers to survey questions.(11)

Physicians in primary care spend more time documenting care than other physicians, so it is vital to have high quality data sources about how they use EHRs.(12) In particular, it is important to find policies that maximize the benefits of EHRs while minimizing their potential to add to physicians’ burdens.(13,14) Our findings suggest that data collected by certifying boards, such as ABFM, have several appealing characteristics that could make them useful as a supplement to NEHRS. First, the data are likely to have a higher assurance of representativeness and reliability, especially if collected as a mandatory portion of recertification activities. Next, these certifying boards could collect data that are more focused on the EHR experience of physicians within particular medical specialties. Certifying boards could also leverage their relationships with physicians to collect ongoing qualitative information about emerging issues with EHRs and other medical technologies that can inform future surveys. However, individual certifying boards will by definition only be able to represent the views of an individual primary specialty and any related subspecialties.

### Limitations

There were several limitations to this work. Despite coordination between the three institutions, there were some differences in the questions or their response options. Some questions, such as the question about practice size, had been carried forward from previous versions of the CCQ. This limited our ability to reach more detailed conclusions about some potential comparisons. Perhaps more substantially, the CCQ only included physicians within one subspecialty of primary care, albeit the third largest medical specialty. There are some differences between family physicians and other types of primary care physicians, including that family physicians are more likely to practice in rural and underserved areas.(15) In many other ways, though, family physicians are quite similar to other physicians working in primary care.(16,17) Without information from other primary care specialties, it is impossible to know what impact this has on our conclusions.

## Conclusion

Our study compared the NEHRS, a key national survey used to inform policymakers on the state of HIT, with other surveys and found that while similar conclusions could be drawn at a high level related to the state of interoperability and physicians’ experiences documenting in EHRs, there were also significant differences when looking at the findings at a more granular level. There were also important differences in the composition of the respondents which likely impacts the results of these surveys. These differences are meaningful for policymakers whose objectives are to advance the development and use of HIT and improving data sharing in service to patient care. Future work is needed to assess variation in self-reported HIT experiences of physicians in other medical subspecialties. The strategy of augmenting NEHRS with data from the recertification surveys of other certifying boards would be extremely valuable for the high reliability they offer.

## Funding

Office of the National Coordinator for Health Information Technology, Department of Health and Human Services, Cooperative Agreement Grant # 90AX0032/01-02

## Acknowledgments

We gratefully thank the respondents of all surveys for their participation.

## Supplemental material for “Comparative Analysis of Three Surveys on Primary Care Providers’ Experiences with Interoperability in Electronic Health Records”

### I. Documentation and visit practices

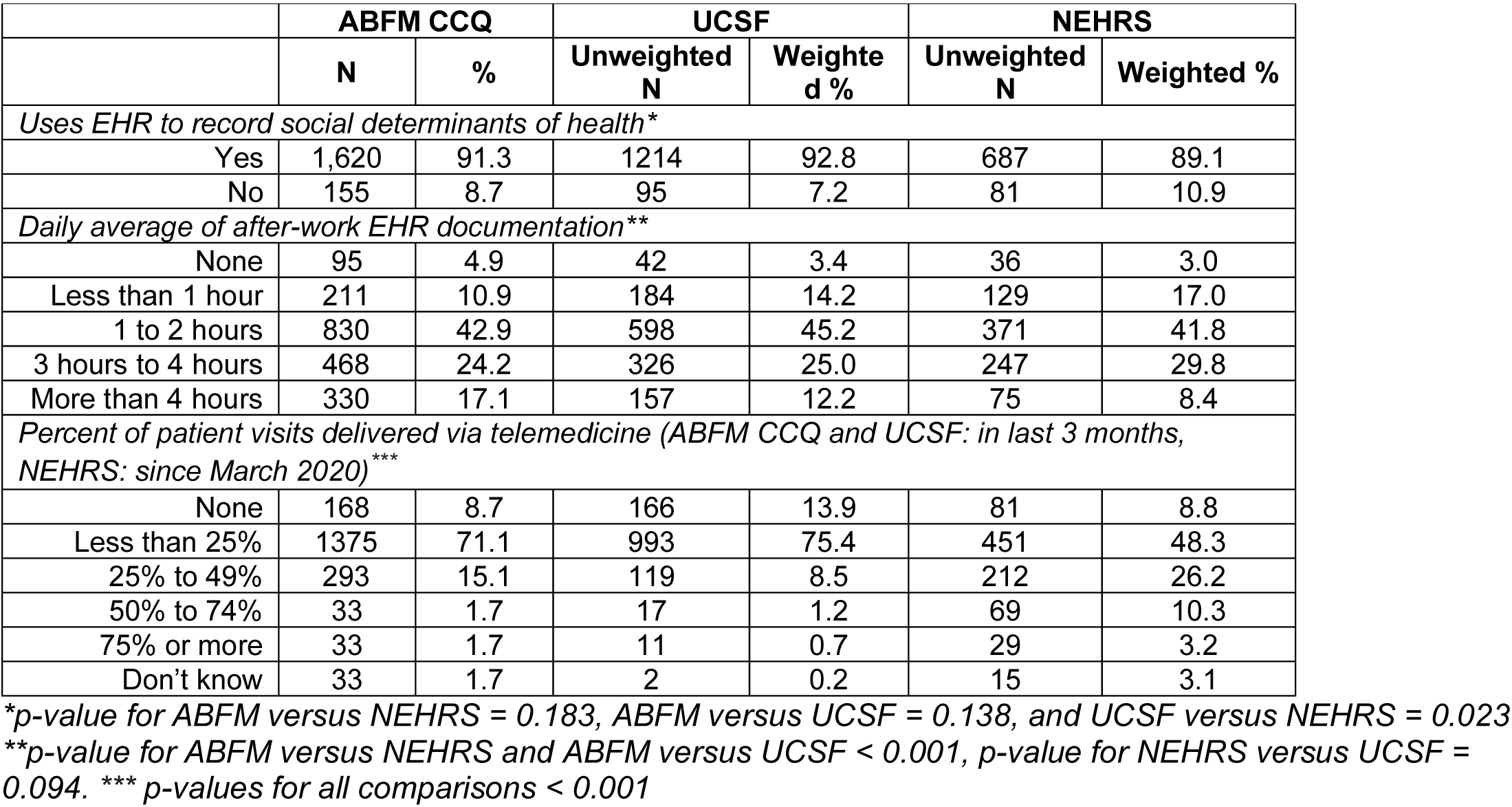

|  | **ABFM CCQ** |  | **UCSF** |  | **NEHRS** |  |
| --- | --- | --- | --- | --- | --- | --- |
|  | **N** | **%** | **Unweighted N** | **Weighted %** | **Unweighted N** | **Weighted %** |
| *Uses EHR to record social determinants of health\** |  |  |  |  |  |  |
| Yes | 1,620 | 91.3 | 1214 | 92.8 | 687 | 89.1 |
| No | 155 | 8.7 | 95 | 7.2 | 81 | 10.9 |
| *Daily average of after-work EHR documentation\*\** |  |  |  |  |  |  |
| None | 95 | 4.9 | 42 | 3.4 | 36 | 3.0 |
| Less than 1 hour | 211 | 10.9 | 184 | 14.2 | 129 | 17.0 |
| 1 to 2 hours | 830 | 42.9 | 598 | 45.2 | 371 | 41.8 |
| 3 hours to 4 hours | 468 | 24.2 | 326 | 25.0 | 247 | 29.8 |
| More than 4 hours | 330 | 17.1 | 157 | 12.2 | 75 | 8.4 |
| *Percent of patient visits delivered via telemedicine (ABFM CCQ and UCSF: in last 3 months, NEHRS: since March 2020)\*\*\** |  |  |  |  |  |  |
| None | 168 | 8.7 | 166 | 13.9 | 81 | 8.8 |
| Less than 25% | 1375 | 71.1 | 993 | 75.4 | 451 | 48.3 |
| 25% to 49% | 293 | 15.1 | 119 | 8.5 | 212 | 26.2 |
| 50% to 74% | 33 | 1.7 | 17 | 1.2 | 69 | 10.3 |
| 75% or more | 33 | 1.7 | 11 | 0.7 | 29 | 3.2 |
| Don't know | 33 | 1.7 | 2 | 0.2 | 15 | 3.1 |

\*p-value for ABFM versus NEHRS = 0.183, ABFM versus UCSF = 0.138, and UCSF versus NEHRS = 0.023
\*\*p-value for ABFM versus NEHRS and ABFM versus UCSF < 0.001, p-value for NEHRS versus UCSF = 0.094. \*\*\* p-values for all comparisons < 0.001

### II. Integration into care of patient information from external organizations

|  | ABFM |  | UCSF |  |
| --- | --- | --- | --- | --- |
|  | N | % | Unweighted N | Weighted % |
| *When you access clinical information from outside your organization (e.g. referrals, consult notes, discharge summaries, patient records) through any means (e.g. fax, phone, EHR, etc), how easy is it to use the information to effectively care for your patients?\** |  |  |  |  |
| Not at all | 158 | 8.2 | 122 | 18.5 |
| Somewhat | 1270 | 65.9 | 838 | 62.8 |
| Very | 465 | 24.1 | 241 | 18.6 |
| Don't Know | 33 | 1.7 | 2 | 0.1 |
| *When you access clinical information about your patients from outside your organization (e.g. referrals, consult notes, discharge summaries, patient records), how often is it from within your EHR in any integrated format (as opposed to a PDF)?\** |  |  |  |  |
| Never | 299 | 15.3 | 249 | 23.9 |
| Rarely | 256 | 13.1 | 217 | 17.8 |
| Sometimes | 630 | 32.3 | 398 | 28.1 |
| Often | 618 | 31.7 | 360 | 24.4 |
| Don't Know | 145 | 7.4 | 75 | 5.7 |
| *How easy is it to use clinical information from clinicians outside your organization that use the same EHR Vendor?\** |  |  |  |  |
| Not at all | 250 | 13.8 | 179 | 19.1 |
| Somewhat | 646 | 35.6 | 459 | 36.3 |
| Very | 765 | 42.2 | 500 | 35.6 |
| Don't Know | 152 | 8.4 | 91 | 9.1 |
| *How easy is it to use clinical information from clinicians outside your organization that use a different EHR Vendor?\** |  |  |  |  |
| Not at all | 615 | 32.6 | 726 | 54.3 |
| Somewhat | 947 | 50.3 | 455 | 36.8 |
| Very | 150 | 8.0 | 41 | 4.0 |
| Don't Know | 172 | 9.1 | 66 | 4.9 |
| *When looking for or using clinical information from outside your organization, to what extent do the following occur: Difficulty finding important information due to a large amount of low-value information\** |  |  |  |  |
| Not at all | 99 | 5.4 | 25 | 2.1 |
| To Some Extent | 883 | 48.1 | 330 | 24.4 |
| To a Great Extent | 855 | 46.5 | 932 | 73.5 |

\*p-value < 0.001

### III. Stratified comparisons

| **EHR Platform** | **Epic** |  |  | **Other Platform** |  |  |
| --- | --- | --- | --- | --- | --- | --- |
|  | **ABFM** | **NEHRS** | **UCSF** | **ABFM** | **NEHRS** | **UCSF** |
|  | % (95% CI) | % (95% CI) | % (95% CI) | % (95% CI) | % (95% CI) | % (95% CI) |
| Very satisfied with EHR | 37 (34 – 39) | 42 (31-54) | 25 (22-29) | 20 (18 – 22) | 24 (19-30) | 16 (13-19) |
| More than 4 hours daily after-work documentation | 17 (15 – 19) | 11 (6-21) | 12 (9-14) | 17 (15 – 19) | 7 (5-11) | 13 (10-16) |
| Integrates patient information | 59 (53 – 65) | 78 (63-88) | 45 (41-49) | 37 (32 – 41) | 47 (40-54) | 32 (28-36) |
| Information from outside encounters sometimes/often available | 83 (79 – 87) | 84 (71-92) | 78 (75-81) | 47 (42 – 52) | 56 (49-62) | 31 (28-36) |
| **Practice Site** | **Private/Independent Practice** |  |  | **Other** |  |  |
| Very satisfied with EHR | 25 (23 – 28) | 29 (23-36) | 23 (19-27) | 27 (26 – 29) | 23 (20-40) | 17 (14-20) |
| More than 4 hours daily after-work documentation | 17 (14 – 20) | 8 (6-12) | 13 (10-17) | 17 (15 – 19) | 9 (4-18) | 11 (9-14) |
| Integrates patient information | 43 (36 – 49) | 50 (43-58) | 38 (33-43) | 47 (43 – 52) | 67 (56-76) | 37 (33-40) |
| Information from outside encounters sometimes/often available | 52 (46 – 59) | 62 (55-68) | 44 (39-49) | 66 (62 – 70) | 71 (60-80) | 55 (51-59) |
| **Age** | **<50** |  |  | **50+** |  |  |
| Very satisfied with EHR | 29 (27 – 30) | 32 (24-43) | 21 (17-26) | 24 (22 – 26) | 27 (21-34) | 19 (16-22) |
| More than 4 hours daily after-work documentation | 15 (13 – 17) | 10 (5-18) | 11 (8-15) | 20 (17 – 23) | 8 (5-11) | 13 (11-16) |
| Integrates patient information | 47 (42 – 52) | 60 (49-70) | 41 (35-47) | 44 (39 – 50) | 52 (45-60) | 36 (33-40) |
| Information from outside encounters sometimes/often available | 63 (59 – 68) | 67 (58-76) | 53 (47-59) | 60 (54 – 65) | 62 (55-69) | 50 (46-53) |

## Notes

### Competing Interest Statement

The authors have declared no competing interest.

### Funding Statement

This research was funded by the United States Office of the National Coordinator for Health Information Technology, Department of Health and Human Services, Cooperative Agreement Grant # 90AX0032/01-02

### Author Declarations

IRB of American Academy of Family Practice waived ethical approval for this work. IRB of University of California San Francisco waived ethical approval for this work.

## References

1. NEHRS - National Electronic Health Records Survey Homepage [Internet]. 2022 [cited 2022 Jul 26]. Available from: https://www.cdc.gov/nchs/nehrs/about.htm

2. Pylypchuk Y, Everson J, Charles D, Patel V. Interoperability Among Office-Based Physicians in 2015, 2017, and 2019. Washington, D.C.: Office of the National Coordinator for Health Information Technology; 2022 Feb. (ONC Data Brief). Report No.: 59.

3. Tripathi M, Yeager M. TEFCA Live! The Future Of Network Interoperability Is Here [Internet]. 2023 [cited 2023 Dec 19]. Available from: http://www.healthaffairs.org/do/10.1377/forefront.20231211.267976/full/

4. 2021 NEHRS public use file documentation [Internet]. 2022 [cited 2022 Jul 26]. 2021 NEHRS public use file documentation. Available from: https://www.cdc.gov/nchs/data/nehrs/NEHRS2021Doc-508.pdf

5. Meyer VM, Benjamens S, Moumni ME, Lange JFM, Pol RA. Global Overview of Response Rates in Patient and Health Care Professional Surveys in Surgery. Ann Surg. 2022 Jan;275(1):e75–81.

6. Sharma AE, Knox M, Peterson LE, Willard-Grace R, Grumbach K, Potter MB. How Is Family Medicine Engaging Patients at the Practice-Level?: A National Sample of Family Physicians. J Am Board Fam Med. 2018 Sep 1;31(5):733–42.

7. Rao JN, Scott AJ. A simple method for the analysis of clustered binary data. Biometrics. 1992 Jun;48(2):577–85.

8. R Core Team. R: A language and environment for statistical computing. R Foundation for Statistical Computing. Vienna, Austria; 2021.

9. Lumley T. Analysis of Complex Survey Samples. J Stat Soft [Internet]. 2004 [cited 2022 Jul 27];9(8). Available from: http://www.jstatsoft.org/v09/i08/

10. Office of the National Coordinator for Health Information Technology. NEHRS Dataset documentation [Internet]. [cited 2022 Aug 8]. Available from: https://ftp.cdc.gov/pub/Health_Statistics/NCHS/Dataset_Documentation/NEHRS/

11. Krosnick JA. Response strategies for coping with the cognitive demands of attitude measures in surveys. Applied Cognitive Psychology. 1991;5(3):213–36.

12. Rotenstein LS, Holmgren AJ, Downing NL, Bates DW. Differences in Total and After-hours Electronic Health Record Time Across Ambulatory Specialties. JAMA Intern Med. 2021 Jun;181(6):863–5.

13. Arndt BG, Beasley JW, Watkinson MD, Temte JL, Tuan WJ, Sinsky CA, et al. Tethered to the EHR: Primary Care Physician Workload Assessment Using EHR Event Log Data and Time-Motion Observations. The Annals of Family Medicine. 2017 Sep 1;15(5):419–26.

14. Robertson SL, Robinson MD, Reid A. Electronic Health Record Effects on Work-Life Balance and Burnout Within the I3 Population Collaborative. Journal of Graduate Medical Education. 2017 Aug 1;9(4):479–84.

15. Talib Z, Jewers MM, Strasser JH, Popiel DK, Goldberg DG, Chen C, et al. Primary Care Residents in Teaching Health Centers: Their Intentions to Practice in Underserved Settings After Residency Training. Academic Medicine. 2018 Jan;93(1):98–103.

16. Xierali IM, Nivet MA. The Racial and Ethnic Composition and Distribution of Primary Care Physicians. J Health Care Poor Underserved. 2018;29(1):556–70.

17. Jabbarpour Y, Westfall J. Diversity in the Family Medicine Workforce. Family Medicine. 2021;53(7):640–3.

